# Individual and city-level variations in heat-related road traffic deaths in Latin America

**DOI:** 10.1101/2025.09.05.25334734

**Authors:** Cheng-Kai Hsu, D. Alex Quistberg, Brisa N. Sánchez, Josiah L. Kephart, Usama Bilal, Nelson Gouveia, Carolina Perez Ferrer, Waleska T. Caiaffa, Amélia Augusta de Lima Friche, Ignacio Yannone, Daniel A. Rodríguez

## Abstract

Road-traffic mortality and extreme heat are two major urban health challenges, increasingly found to be associated. However, few studies have examined this association in Latin America—one of the world’s most urbanized, fastest-motorizing region, with a high share of vulnerable road users— and even fewer have analyzed multiple cities across diverse climates and urban settings. Using temperature and road-traffic mortality data (2000–2019) from 272 cities in six Latin American countries, we conducted a time-stratified case-crossover study. The relative risks (RRs) of road-traffic mortality at the 95th and 99th temperature percentiles, compared to the minimum mortality temperature percentile, were 1.16 [95% CI: 1.14–1.19] and 1.18 [1.15–1.21], respectively. Risks were particularly high among adolescents, males, motorcyclists, bicyclists, and in cities with hotter climates and longer commutes. Policymakers in the tropical Global South should prioritize protecting vulnerable road users in peripheral communities, where many endure long, heat-exposed commutes in non-climate-controlled informal transport.

## Main

The adverse impacts of ambient heat on human health are increasingly documented and expected to intensify given global warming, especially in urban areas. Through the urban heat island effect, global rapid urbanization has amplified human exposure to extreme heat ^1,2^, leading to increased mortality risks across different regions ^3,4^, including unintentional injuries ^5^. Recently, studies have linked road-traffic injuries—responsible for 1.25 million deaths worldwide annually ^6^—to high temperatures ^7^, likely driven by a combination of factors: prolonged heat exposure on roadways; vehicle malfunctions ^8^; deteriorating road conditions like softened asphalt ^9^; increased traffic volume ^10^; strained emergency medical services due to higher call volumes and delayed response times ^11^; and safety-related behavioral changes, such as impaired focus and riskier driving ^12–15^. This phenomenon is critical as its impact may intensify in densely urbanized areas, where there is increased exposure to heat due to the urban heat island effect, along with increased vulnerability to its effects due to the high concentration and diverse mix of vulnerable road users, such as motorcyclists, bicyclists, and pedestrians, susceptible to immediate heat exposure and physical injuries due to a lack of protective enclosures.

However, evidence on temperature-related road-traffic mortality in Latin America is lacking. Most existing studies have focused on single cities in colder regions, including North America, Europe, and the Western Pacific ^7^. This narrow focus presents several issues. First, temperature-related road-traffic mortality remains understudied in regions with diverse climates and urban conditions, despite their importance in understanding how climate impacts human health risks, including road safety, across different geographic contexts. Second, while global warming has improved winter road conditions and safety in colder regions ^16^, prolonged heat exposure in warmer climates may heighten risks; evidence from the Global Burden of Disease (GBD) study 2019 underscores this concern, showing that heat-related road-traffic mortality is high in tropical regions, especially low- and middle-income countries (LMICs) ^17^. Third, unlike high-income countries where motor vehicles may dominate the road, countries in regions like Latin America have a more diverse travel mode mix, with a greater reliance on motorcycles, bicycles, walking, and public transit, a distinct transportation landscape that may alter the associations observed in other settings.

If applicable, these impacts would be particularly concerning for Latin America, one of the world’s most urbanized regions with over 80% of the population living in cities ^18^. The region’s dense urbanization has intensified heat exposure through the urban heat island effect, with projections indicating rising temperatures and more frequent extreme heat events ^19^. For instance, in South America’s major cities, the number of extremely hot days (defined by the 95th percentile daily mean temperature from 1961 to 1990) is projected to increase five-to ten-fold from the late 20th century to the mid-21st century under the mid-level representative concentration pathway (RCP) 4.5 climate scenario ^20^. Meanwhile, road-traffic mortality has become the leading cause of death for children (5–14 years), and the second leading cause for people aged 15–44 years ^21^, likely driven by urban expansion and rapid motorization. The region’s motorcycle-oriented motorization has furthered these safety concerns ^22,23^, manifesting in a sharp rise in motorcycle mortality rates from 0.8 per 100,000 in 1998 to 3.5 per 100,000 in 2010 ^24^.

This study fills existing research gaps by examining the temperature effects on road-traffic mortality across Latin America. We leverage a valuably large dataset spanning two decades and 272 Latin American cities in six countries from Mexico to the Southern Cone. To our knowledge, this study is the first to explore these effects across multiple cities, offering two strengths: firstly, examining temperature conditions as a risk factor for road-traffic mortality, a relatively rare death event requiring pooling a large dataset for sufficient statistical power; and secondly, exploring variations across demographic groups, transportation modes and cities with varying climatic patterns, transportation settings, and urban design factors. These insights are essential for understanding temperature-related road-traffic mortality, a growing global burden on human health, particularly in rapidly urbanizing regions that are warming, motorizing, and increasingly vulnerable to climate change impacts.

## Results

### Data description

We examined 272 cities across six Latin American countries from 2000 to 2019, totaling 1,956,856 city-days. Our study covered Brazil (152 cities, 2000–2019), Mexico (92 cities, 2000–2019), Chile (21 cities, 2000–2019), El Salvador (three cities, 2001–2019), Panama (three cities, 2002–2019), and Costa Rica (one city, 2000–2019). Table 1 presents summary statistics for road-traffic mortality, stratified by age, binary sex, and mode of transportation involved in the fatality, alongside temperature conditions. Mortality statistics were based on the median values of a multiply imputed dataset (across 100 imputations), which addressed ill-defined or partially classified ICD-10 (International Classification of Diseases, 10th Revision) codes (see Methods for details). The mortality analysis followed two steps: first, calculating the daily average number of road-traffic deaths for each city (scaled to deaths per 100 days), and second, summarizing these averages across 272 cities. On average, there were 33.75 road-traffic deaths per 100 days (SD: 72.52, Median: 14.80, Range: 3.42–717.06). By mode, deaths per 100 days averaged 7.50 for motorists, 5.09 for motorcyclists, 0.90 for bicyclists, and 13.87 for pedestrians. By sex, the average was 26.81 for males and 6.93 for females. Age-stratified averages were 1.11 (<9 years), 3.27 (10– 19), 11.56 (20–34), 13.58 (35–64), and 4.22 (≥65). Mean daily temperature was 21.20°C (SD: 3.99°C).

**Table 1.** Descriptive statistics of study variables using city-level averages per 100 days.

|  | Mean | SD | Median | Min | Max |
| --- | --- | --- | --- | --- | --- |
| *Outcome (Number of Road-traffic Deaths per 100 days)* |  |  |  |  |  |
| Overall | 33.75 | 72.52 | 14.80 | 3.42 | 717.06 |
| <u>By Mode of Transportation</u> |  |  |  |  |  |
| Automobile | 7.50 | 15.93 | 3.64 | 0.00 | 194.50 |
| Motorcycle | 5.09 | 12.21 | 1.88 | 0.00 | 151.66 |
| Bicycle | 0.90 | 1.66 | 0.37 | 0.00 | 16.00 |
| Pedestrian | 13.87 | 35.80 | 5.30 | 0.03 | 367.45 |
| <u>By Sex</u> |  |  |  |  |  |
| Male | 26.81 | 56.89 | 11.90 | 2.59 | 578.01 |
| Female | 6.93 | 15.79 | 2.94 | 0.70 | 168.24 |
| <u>By Age</u> |  |  |  |  |  |
| ≤9 | 1.11 | 2.31 | 0.49 | 0.04 | 24.68 |
| 10-19 | 3.27 | 7.28 | 1.46 | 0.19 | 78.57 |
| 20-34 | 11.56 | 24.65 | 5.18 | 0.93 | 255.95 |
| 35-64 | 13.58 | 28.58 | 5.83 | 1.39 | 282.44 |
| ≥65 | 4.22 | 10.25 | 1.68 | 0.10 | 111.78 |
| *Exposure* |  |  |  |  |  |
| Average Daily Temperature (°C) | 21.20 | 3.99 | 21.55 | 5.17 | 28.16 |
| Average Daily Temperature (%tile) | 49.85 | 0.54 | 49.82 | 48.33 | 54.12 |

### Associations between temperature conditions and road-traffic mortality

This section outlines key findings. We first describe the main effects analysis, which considered overall road mortality as the outcome and pooled estimates across all cities. Next, we present the stratification analysis by temperature cluster, assessing how temperature-mortality associations vary across groups of cities with distinct temperature ranges. This is followed by the subgroup analysis, which stratified the outcome by victims’ sex, age, and mode of transportation involved in the mortality. Lastly, we present the effect modification analysis, which investigated interactions between temperature conditions and two city-level effect modifiers: mean street-segment length (km) and mean peak-hour travel time (minutes). Table 2 summarizes the results of the main effects, stratification, and subgroup analyses using two key metrics: relative risk (RR) and excess death fraction (EDF). The RR quantifies the likelihood of road-traffic mortality at the 95th and 99th temperature percentiles relative to the minimum mortality temperature percentile (MMTP), defined as the temperature percentile with the lowest RR between the 1st and 99th percentiles in the main analysis. The EDF represents the proportion of total road-traffic mortality attributable to extreme heat (≥95th percentile).

**Table 2.** **Estimated RRs at the 95^th^ and 99^th^ temperature percentile (relative to the minimum mortality temperature percentile) and estimated extreme-heat related EDFs for the main effects, stratification, and subgroup analyses**.

|  | RR at 95<sup>th</sup> %tile | RR at 99<sup>th</sup> %tile | EDFs from extreme heat (>95<sup>th</sup> %tile) |
| --- | --- | --- | --- |
| Overall | 1.16 [95%CI: 1.14, 1.19] | 1.18 [95%CI: 1.15, 1.21] | 0.74% [95%eCI: 0.64%, 0.84%] |
| <u>By temperature cluster</u> |  |  |  |
| Hot and Dispersed | 1.34 [1.21, 1.48] | 1.42 [1.27, 1.59] | 1.46% [1.01%, 1.87%] |
| Hot and Concentrated | 1.16 [1.12, 1.21] | 1.16 [1.11, 1.22] | 0.71% [0.53%, 0.88%] |
| Warm and Dispersed | 1.18 [1.10, 1.26] | 1.21 [1.12, 1.31] | 0.84% [0.49%, 1.15%] |
| Warm and Concentrated | 1.17 [1.13, 1.21] | 1.21 [1.15, 1.26] | 0.81% [0.62%, 0.98%] |
| Mild | 1.10 [1.00, 1.20] | 1.09 [0.98, 1.21] | 0.44% [-0.06%, 0.87%] |
| Cool | 1.08 [1.01, 1.15] | 1.07 [0.99, 1.17] | 0.32% [-0.04%, 0.59%] |
| <u>By sex</u> |  |  |  |
| Male | 1.17 [1.15, 1.20] | 1.19 [1.16, 1.23] | 0.79% [0.68%, 0.90%] |
| Female | 1.12 [1.07, 1.17] | 1.13 [1.07, 1.19] | 0.55% [0.30%, 0.77%] |
| <u>By age</u> |  |  |  |
| ≤9 years | 1.24 [1.10, 1.39] | 1.25 [1.10, 1.43] | 0.99% [0.46%, 1.49%] |
| 10-19 years | 1.26 [1.18, 1.35] | 1.26 [1.16, 1.37] | 1.06% [0.74%, 1.36%] |
| 20-34 years | 1.16 [1.12, 1.21] | 1.17 [1.12, 1.22] | 0.73% [0.55%, 0.90%] |
| 35-64 years | 1.17 [1.13, 1.21] | 1.20 [1.15, 1.24] | 0.79% [0.63%, 0.95%] |
| ≥ 65 years | 1.06 [1.00, 1.12] | 1.08 [1.02, 1.15] | 0.32% [0.04%, 0.59%] |
| <u>By mode</u> |  |  |  |
| Motorcycle | 1.24 [1.18, 1.30] | 1.27 [1.20, 1.34] | 1.17% [0.93%, 1.42%] |
| Bicycle | 1.21 [1.09, 1.34] | 1.21 [1.07, 1.36] | 0.88% [0.40%, 1.36%] |
| Pedestrian | 1.14 [1.10, 1.17] | 1.16 [1.11, 1.20] | 0.60% [0.47%, 0.74%] |
| Motor vehicle | 1.14 [1.10, 1.19] | 1.15 [1.10, 1.21] | 0.66% [0.46%, 0.87%] |

Fig. 1 presents the main effects analysis, showing the estimated association between temperature conditions and road-traffic mortality, derived by pooling data across all 272 cities. When based on temperature percentiles, the main effects model shows a monotonic increasing trend in the estimated association curve (Fig. 1a). We observed RRs of 1.16 [1.14–1.19] and 1.18 [1.15–1.21] at the 95th and 99th percentiles compared to the MMTP, respectively (Table 2). The EDF for temperatures above the 95^th^ percentile indicates that 0.74% [0.64%–0.84%] of road-traffic mortality was attributable to extreme heat (Table 2). When based on absolute temperatures in °C, the curve appears to taper off at the lowest and highest ends of the temperature (Fig. 1b), with wide confidence intervals (CIs) reflecting the relatively fewer observations in these ranges (Fig. 1d). While the curve in Fig. 1b shows some non-linearity, it remains similar to that in Fig. 1a within the 1st and 99th percentiles (marked in red within the leftmost and rightmost dashed lines). In sensitivity analyses, varying the knot specifications yielded highly similar results (Supplementary Fig. 1).

**Fig. 1.**
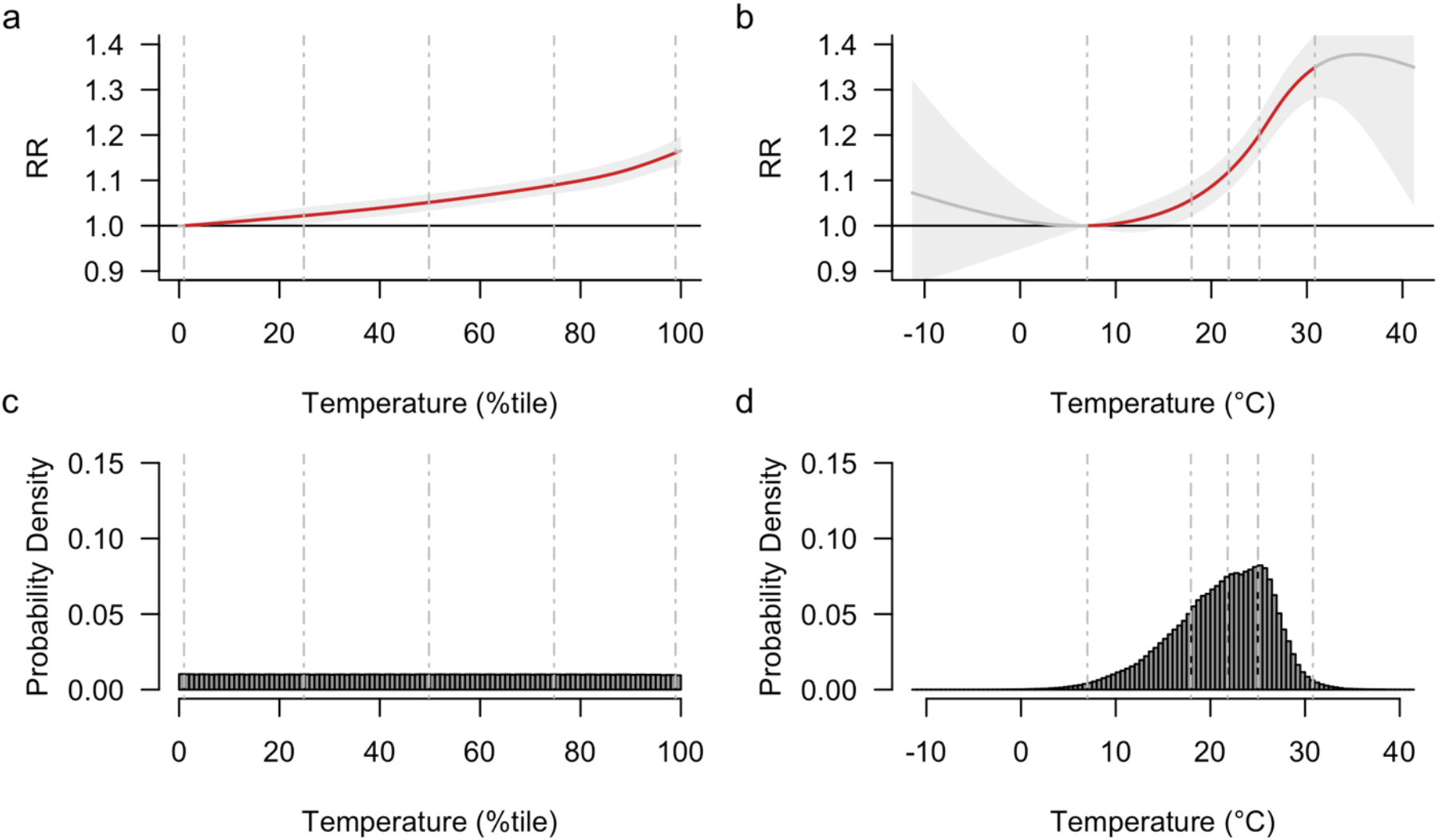
Association between temperature conditions and road-traffic mortality (main effects model). (a) Relative temperature measure (temperature percentiles). (b) Absolute temperature measure (°C). (c) Distribution of temperature percentiles. (d) Distribution of absolute temperatures (°C). Dashed vertical lines indicate temperature percentiles: 1^st^, 25^th^, 50^th^, 75^th^, and 99^th^ from left to right. Shading in each association curve represents the 95% confidence interval.

Fig. 2 displays the stratification analysis by temperature cluster. Each temperature cluster was labeled according to the overall central tendency and spread of the temperature distributions for the cities in the cluster (Fig. 2a). For example, the “Cool” cluster included cities with an average temperature of 12.6°C (1^st^–99^th^ percentile: 1.6–19.6°C), whereas the “Hot & Dispersed” cluster had the highest average temperature (23.0°C) and a broader range (1^st^–99^th^ percentile: 6.4–35.2°C). Figs. 2b–g illustrate the estimated effects for each cluster. Except for the Cool Cluster and Mild Cluster, the remaining four temperature clusters generally demonstrated significant associations, with the Hot & Dispersed Cluster exhibiting the most pronounced effect (Fig. 2g), with estimated RRs of 1.34 [1.21–1.48] and 1.42 [1.27–1.59] at the 95th and 99th percentiles, and an extreme heat-related EDF of 1.46% [1.01%–1.87%] (see Table 2 for RRs for each temperature cluster).

**Fig. 2.**
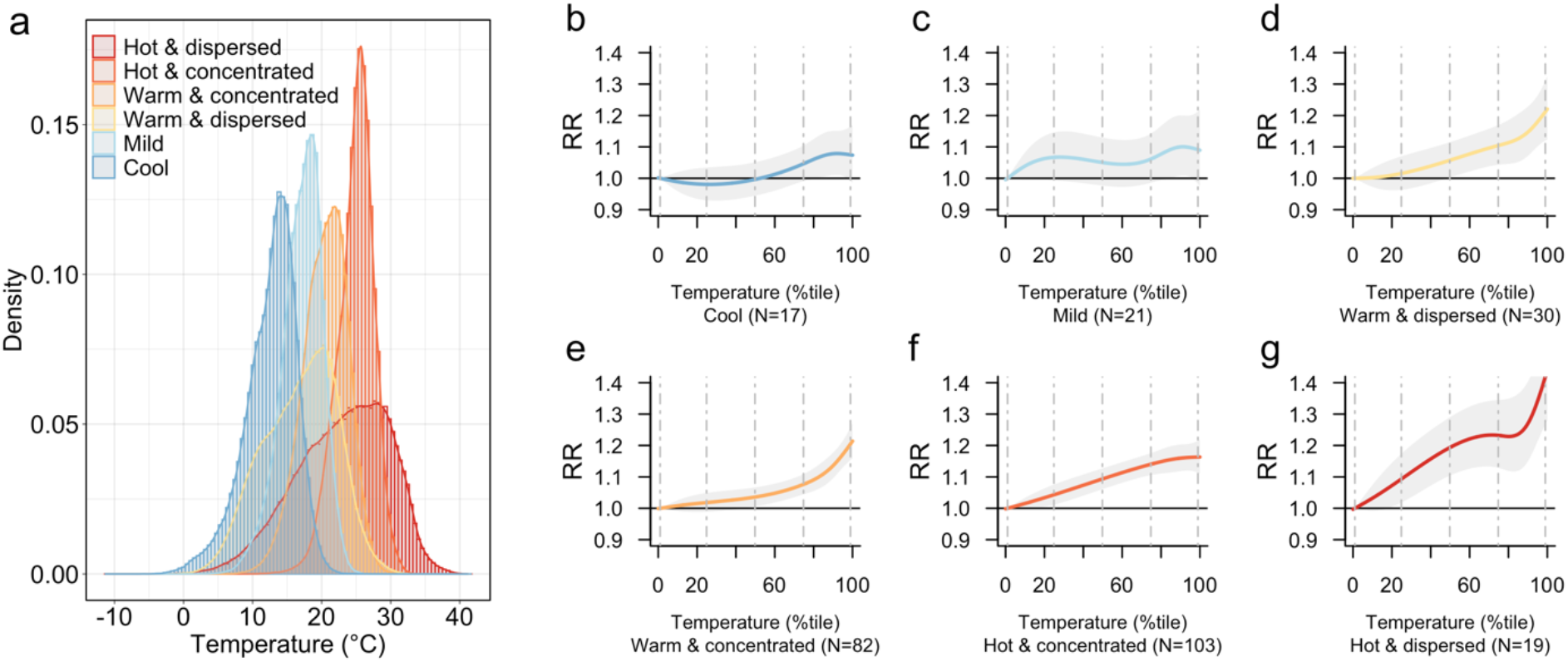
Associations between temperature and road-traffic mortality by temperature cluster. (a)Temperature distribution (°C) for each cluster. (b)-(g) Association between temperature percentiles (x-axis) and road-traffic mortality for each temperature cluster. N indicates number of cities in each cluster. Dashed vertical lines in figs. b through g indicate temperature percentiles: 1^st^, 25^th^, 50^th^, 75^th^, and 99^th^ from left to right. Shading in each association curve represents the 95% confidence interval.

We conducted sensitivity analyses assessing whether city-level temperature characteristics—specifically, average temperature (°C) and temperature standard deviation (°C)— modified the association. Tests of effect modification indicated significantly stronger temperature-mortality associations in cities with higher average temperatures and greater temperature variability (Supplementary Fig. 2). Specifically, Supplementary Fig. 2a shows that cities with higher average temperatures were generally more susceptible to heat-related road mortality risks compared to colder cities, as indicated by a Chi-square likelihood ratio test (χ^2^(12) = 26.848, p = 0.018). Similarly, Supplementary Fig. 2b shows that cities with more varied temperature distributions (higher standard deviations) faced an even greater susceptibility to heat-induced road mortality risks (χ^2^(12) = 57.582, p < 0.001), with risks increasing sharply around the 95th percentile. While pooling data across all cities merges temperature distributions with varying means and standard deviations—potentially masking temperature cluster-specific patterns—the findings in Fig. 2 and Supplementary Fig. 2 consistently reveal an increased risk of road-traffic mortality with rising temperatures. The magnitude of this risk varied according to the temperature distribution, whether analyzed by temperature cluster or by mean and standard deviation, with the highest risks observed in the hottest cities and those experiencing greater temperature variability.

Fig. 3 presents the results of subgroup analyses by victim demographics and transportation mode involved in the fatality. We observed that males consistently had higher RRs than females (RRs at 99th percentile: males, 1.19 [1.16, 1.23]; females, 1.13 [1.07, 1.19]; Table 2), with a sharp increase beyond the 80^th^ percentile (Fig. 3a). Children (<9 years) and adolescents (10-19 years) had higher vulnerability than other age-groups, with adolescents being particularly vulnerable (steepest association curve, Fig. 3b). Young (20-34 years) and middle-aged adults (35-64 years) showed intermediate vulnerability while older adults (≥65 years) exhibited the lowest vulnerability. Motorcyclists and bicyclists had higher RRs than motor vehicle users and pedestrians (Fig. 3c), with motorcycle mortality increasing sharply after the 80^th^ percentile and reaching an RR of 1.27 [1.20–1.34] at the 99^th^ percentile (Table 2).

**Fig. 3.**
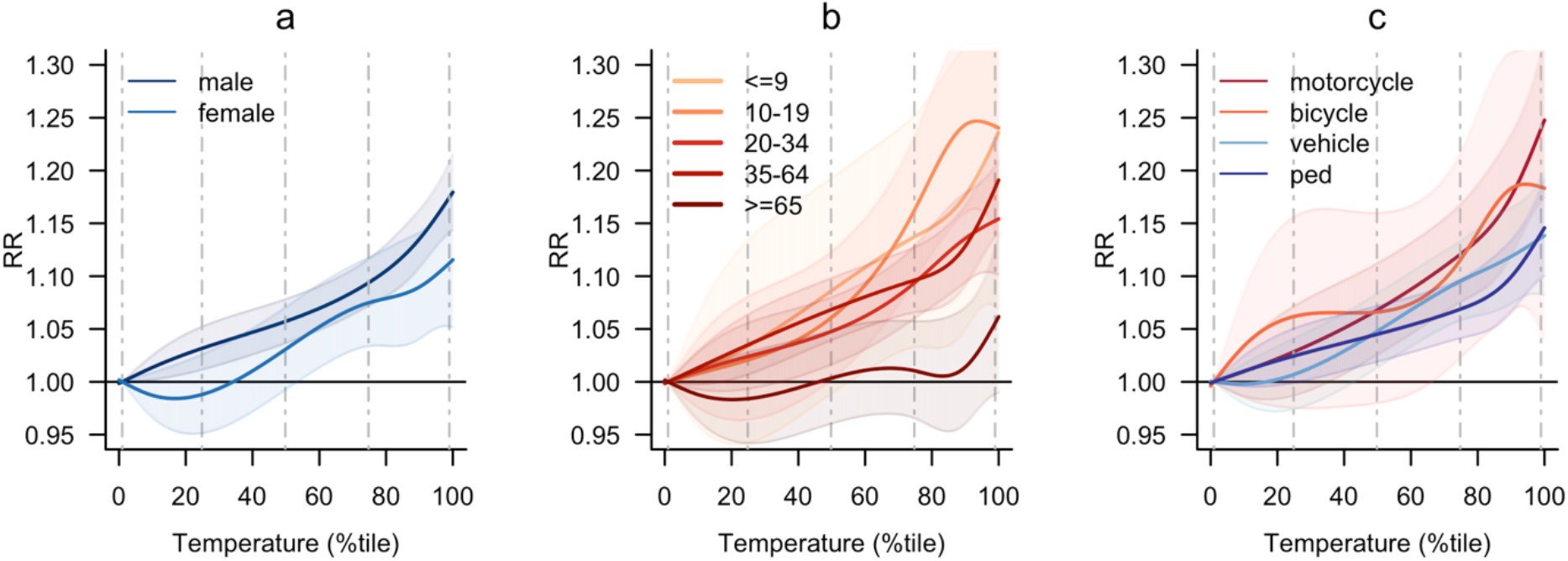
Sub-group analysis results. Relative risk of road mortality at each temperature percentile in reference to the 1^st^ temperature percentile. (a) Sex. (b) Age. (c) Mode of transportation. Dashed vertical lines indicate temperature percentiles: 1^st^, 25^th^, 50^th^, 75^th^, and 99^th^ from left to right. Shading in each association curve represents the 95% confidence interval.

Fig. 4 shows effect modification analysis by city characteristics. We found that residents of cities with longer average street-segments (at the 90^th^ percentile) appeared to exhibit greater vulnerability than those of cities with shorter segments (Chi-square likelihood ratio test, χ^2^(12) = 20.576, p = 0.096; Fig. 4a). Similarly, residents of cities with longer peak-hour travel times showed higher vulnerability than those of cities with shorter peak-hour travel times (χ^2^(12) = 23.048, p = 0.051; Fig. 4b).

**Fig. 4.**
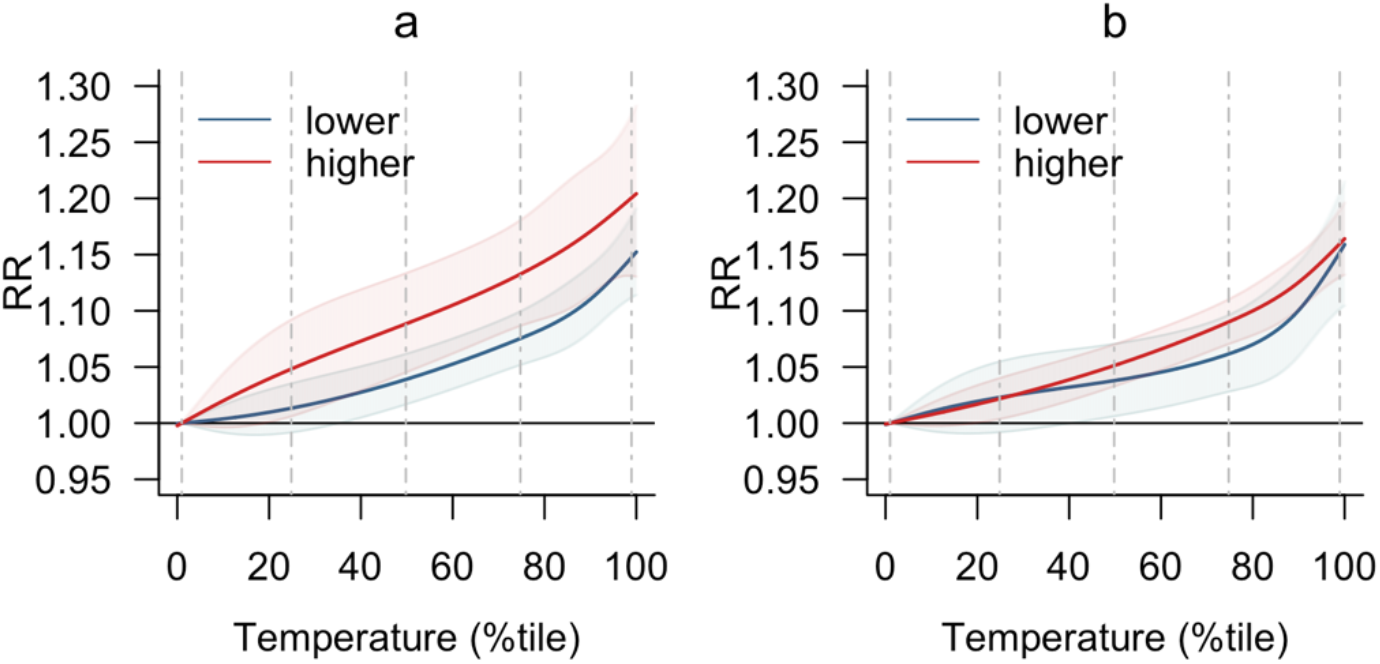
Effect modification from city-level transportation-related factors. (a)Average street-segment length. (b) Average peak-hour travel time. High (red) and low (blue) indicate cities at the 90^th^ percentile (higher values) or 10^th^ percentile (lower values) of respective variable values. Dashed vertical lines indicate temperature percentiles: 1^st^, 25^th^, 50^th^, 75^th^, and 99^th^ from left to right. Shading in each association curve represents the 95% confidence interval.

## Discussion

Analyzing 1,956,856 city-days across 272 cities over two decades, we have identified a distinct association between temperature condition and road-traffic mortality in Latin America, characterized by a monotonic increase, contrasting with the U-shaped curves previously reported in colder locations like Northern China ^7^ and South Korea ^25^. The observed association further contrasts those observed for all-cause, cardiovascular, and respiratory mortality in the region ^4^ and elsewhere ^26,27^. Unlike other causes of mortality influenced by both cold and heat, road-traffic mortality appears primarily heat-driven—at least in our sample, where no city had an average daily temperature below 5 °C during the study period (Table 1). Notably, our estimated extreme heat-related EDF of 0.74% exceeds those reported for all-cause, cardiovascular, and respiratory mortality in similar Latin American contexts using the same extreme heat definition in a SALURBAL study ^4^. This is alarming, as while global warming may reduce certain mortality risks related to cold ^28^, heat-induced road-traffic mortality may continue to rise without such compensatory effects, particularly in the region’s hotter cities, where we observed the strongest effects (Fig. 2g) and the highest EDF (Table 2).

Prior research suggests several causal pathways through which heat may elevate the risk of road-traffic collisions and subsequent fatalities. Heat exposure may impair driver performance due to distraction, fatigue, or diminished cognitive function ^14,15^. Heat can also induce road aggression, impatience, and risky driving like red-light running and speeding ^12,13^. This is supported by our finding of stronger effects in cities with longer street-segments, which are often associated with higher-speed travel ^29^. These heat-induced physiological and behavioral responses are likely compounded by additional challenges on hot days, including tire blowouts ^8^, softened asphalt, increased traffic volume ^10^, and strained emergency medical services due to higher call volumes and delayed response times ^11^.

Our study found particularly higher vulnerability among younger individuals (0-19 years), consistent with prior research ^30^. This contrasts with most heat-related causes of mortality, such as cardiovascular and respiratory diseases, which predominantly affect older adults ^4^. Unlike disease-related deaths, where heat typically worsens pre-existing health conditions or physiological vulnerabilities (e.g. greater demands of thermoregulation in older adults ^31^), rather than initiating disease onset or progression, heat can impact road-traffic mortality more directly, affecting even otherwise healthy individuals and leading to immediate fatalities, greater premature deaths, and more years of life lost.

The stronger associations observed among bicyclists and motorcyclists, as well as among males—who constitute the majority of users for both modes ^24^—also align with previous studies ^17,32^, underscoring several health equity concerns in the region. The higher vulnerability among users of bicycles and motorcycles may stem from their open design exposing users to direct heat, and the greater physical and cognitive demands of riding, such as handgrip, balance, and gait function. This risk is especially concerning for lower-income urban populations with restricted access to climate-controlled mobility options, such as motor vehicles. Moreover, the region’s enormous growth in male motorcyclists, often from lower-income backgrounds, working in on-demand delivery services within the gig economy ^33^ further exacerbates these safety concerns, as they are more prone to risky driving, especially when heat-stressed ^34^.

By showing stronger associations for residents of cities with longer commutes, our study raises global health concerns for LMICs, where residents of peripheral, poor communities often face lengthy travel times, as seen in India, Brazil, and Kenya ^35–37^. In addition to the extended heat exposure owing to long commutes, lower-income populations have limited alternatives for adjusting their travel—such as switching modes, modifying schedules, or working remotely— especially for non-discretionary work commutes that require travel at fixed times regardless of weather conditions. This concern can be further amplified in the tropical Global South, where informal transportation relies heavily on semi-enclosed, non-motorized, and non-air-conditioned modes, such as motorcycle taxis in Brazil and Colombia, cycle rickshaws in India and South Africa, keke (tricycles) in Nigeria, and tuk-tuks in Kenya. While popular among the urban poor because these modes often act as gap fillers ^38^, they offer minimal protection from external hazards such as heat. Given their prevalence and the challenging urban conditions in which they operate— including long, heat-exposed commutes—these factors underscore pressing global health inequities amidst a changing climate.

Strengths of our study include its multi-country, multi-city scope, allowing us to explore variations across diverse climates and urban settings, yielding meaningful global health insights. The multi-city approach using a large dataset across 272 diverse cities also offered robustness of our findings on the impact of heat on road-traffic mortality, in contrast to prior research often limiting analysis to single cities ^7^. Additionally, we provide regional insights currently lacking in the literature, which is critical as Latin America presents unique characteristics; unlike regions studied in existing research, Latin America generally has warmer climates, making it more vulnerable to global warming. The region also has a distinct mode mix, with motorcycle-oriented motorization, where motorcyclists—who tend to come from lower-income backgrounds— experience greater heat exposure and, as our findings show, higher vulnerability to heat-related road-traffic mortality, with an extreme heat-related EDF of 1.17% (Table 2), indicating that 1.17% of motorcycle mortality can be attributed to extreme heat.

Several limitations are noted. We used a relative temperature metric, limiting direct assessment of specific temperature thresholds in °C on road-traffic mortality; however, this approach might be necessary in our multi-city study in a region with heterogeneous climatic patterns ^39^. Additionally, we lacked data on daily traffic volume like vehicle-miles traveled—a common limitation in road safety studies. However, controlling for temporal trends and traffic variations using day-of-week indicators has been shown to yield results consistent with analyses that include traffic volume data ^40^. Further, despite the extensive geographical and temporal scope of our analysis, the CIs for some of our results were wide due to the relatively low counts of road-traffic mortality within each subgroup. Future research can acquire more data, enhancing the robustness of our subgroup comparisons. There may also be mismeasurement of mode of transportation in the subgroup analysis given the existence of ill-defined injuries, which we tried to correct for using a redistribution model.

The monotonic increase in road-traffic mortality with higher temperatures, along with higher vulnerability among motorcyclists, bicyclists, and residents of cities with hotter climates and longer commute times, underscores significant public health concerns. These risks are particularly pronounced for lower-income urban populations who heavily rely on these travel modes and face greater exposure—both to severe injuries in the event of a crash and to higher temperatures in traffic. Our findings further raise critical global health equity concerns, particularly in LMICs in the tropical Global South, where peripheral, low-income communities often endure long commutes and rely on semi-enclosed, non-climate-controlled informal transportation. Further research is needed to understand how climate dynamics impact road safety in these settings. Cities should invest in built environments that promote sustainable transportation while also protecting users of these modes, particularly with increasing global temperatures putting them at higher risk of being road-traffic collision victims.

## Methods

### Study setting

This study was conducted as part of the SALURBAL (Salud Urbana en América Latina, “Urban Health in Latin America”) project, an international research initiative consolidating and harmonizing data on environmental, social, and health data across 371 cities from 11 Latin American countries, with populations above 100,000 as of 2010 ^41^. This is valuably large public health dataset which permits us to have sufficient power to examine road-traffic fatalities in relation to ambient temperature.

In this study, we focused on a subset of 272 cities in six countries, including Brazil (152), Mexico (92), Chile (21), El Salvador (3), Panama (3), and Costa Rica (1)—which have sufficiently detailed mortality records (i.e., more detailed ICD-10 codes to specifically identify road-traffic mortality). The years of analysis were determined by the availability of daily road-traffic mortality data, summarized as follows: Brazil (2000-2019), Mexico (2000-2019), Chile (2000-2019), El Salvador (2001-2019), Panama (2002-2019), and Costa Rica (2000-2019).

### Outcome, exposure, and effect modifiers

The outcome variable was the daily count of road-traffic deaths, defined using ICD-10 codes V01-V89 ^42^. Since 11% of deaths from external causes in countries in SALURBAL were coded with ill-defined or partially-defined ICD-10 codes, e.g., Y10-Y34 or X59, we imputed cause of death as either non-road-traffic deaths (excluded from this study) or road-traffic deaths and its subcategories: pedestrian injuries (V01–V09), cyclist injuries (V10–V19), motorcyclist injuries (V20–V29), and motor vehicle occupant injuries (V30–V79), as previously described elsewhere ^42^. Specifically, we fitted a multinomial model with each subcategory or non-road-traffic deaths as the outcome, using age (grouped into 5-year age-groups), sex, year, and country as predictors. We then predicted probabilities of each death being a road-traffic death subcategory, and conducted multinomial draws from these probabilities to create 100 imputed datasets. For each imputed dataset, a daily time-series of road-traffic death counts was constructed, resulting in 100 separate time-series for each city.

The exposure variable was the daily mean temperature, derived from hourly temperature data from ERA5-Land reanalysis at a native 9-km resolution, population-weighted ^43^. Missing temperature values were imputed using a random forest regression model, integrating resampled ERA5 temperature data ^44^ at a 31-km resolution, along with elevation and aspect (terrain orientation). Kriging spatial interpolation was then employed to model the residuals ^4^. To enable data pooling across cities, we ensured complete overlap in exposure ranges by standardizing temperature data, i.e., converting absolute values (°C) into city-specific temperature percentiles (ranging from 0 to 100) based on all available years.

We also collected data on effect modifiers at the city level, specifically mean street-segment length (km) and mean peak-hour travel time (minutes). These variables were selected to capture different dimensions of urban form and mobility landscape that might influence heat-related road-traffic mortality. Mean street-segment length measures the average length of segments between nodes (i.e., intersections) within each city’s street network, with larger values reflecting longer segments or blocks. In the street network data, sourced from OpenStreetMap, each intersection (where two or more streets meet) was considered as a node and the link connecting each node a street-segment ^41^. Mean peak-hour travel time captures typical commute duration during peak-hours; larger values indicate longer commute times. The metric was derived from the Google Maps Distance Matrix API, using estimated delays for 30 randomly selected origin-destination pairs within the street network at seven different time points during peak-hours on a typical weekday ^45^. Both variables were operationalized as higher or lower than the median across all cities.

### Statistical methods

We employed a time-stratified case-crossover (known also as case time-series) design, pooling the data across cities in a single model ^46^. While multi-city analyses often use a two-step approach to pool results across multiple cities (i.e., pooling city-specific estimates using meta-regression ^46^), the approach is not feasible in the context of road-traffic mortality because city-specific models fail to converge when the total number of road-traffic deaths within a city is low ^47^. Thus, we used a single-step approach ^47^, incorporating city into a four-way strata term (city-year-month-day of the week, e.g., Monday), matching each case city-day to controls within the same city, day of the week, month, and year (1:3 or 1:4 matching, depending on whether the month had four or five Mondays). In other words, the number of road-traffic deaths in a Monday of July of 2018 in Mexico City are compared to the number of road-traffic deaths in all other four Mondays of July of 2018 in Mexico City. This design strengthens the causal interpretation of the findings, enables robust multi-city analysis, and addresses the rarity of road-traffic mortality events.

We operationalized the above approach using a distributed lag non-linear model (DLNM) within a conditional quasi-Poisson modeling framework ^48^. The DLNM characterizes both exposure-response and lag-response relationships via a cross-basis term serving as a bi-dimensional parametrization. We placed three knots at the 10th, 75th, and 90th temperature percentiles ^4,28^. We specified a two-day lag because road-traffic deaths frequently occur within this timeframe following incidents ^49^, which is a shorter lag time than heat-related deaths from other pathways. The RRs of road-traffic mortality associated with various temperature levels were estimated with 95% CIs.

Given that our datasets are multiply imputed, we ran the models for each imputed time series separately and then applied Rubin’s rule to pool the coefficient and variance-covariance estimates from each model based on the 100 imputed datasets, allowing us to reconstruct the pooled estimates from 27,200 time series (100 per city). This step was applied to the main effects analysis and all other analyses discussed in the following subsection. In addition, when calculating the RRs, the association curves were consistently centered around the MMTP identified in the main effects analysis, corresponding to the temperature percentile with the lowest RR between the 1^st^ and 99^th^ percentiles, reflecting the lowest risks for overall road-traffic mortality. The EDF was calculated for extreme heat (≥95^th^ percentile), following the same approach used in Kephart and colleagues ^4^ and described elsewhere ^50^.

### Stratification, subgroup and effect modification analyses

For stratification analysis, we stratified models by six clusters of cities characterized by distinct temperature profiles. Specifically, cities were grouped via hierarchical clustering using Ward’s minimum variance method based on five selected temperature percentiles: the 1^st^, 10^th^, 75^th^, 90^th^, and 99^th^. Unlike clustering based solely on summary statistics like mean and variance, this clustering approach considered the entire temperature distribution. The use of the 1^st^ and 99^th^ percentiles instead of minimum and maximum values attenuated outlier influence. The 10^th^, 75^th^, and 90^th^ percentiles were included as they are commonly used in heat-related mortality research ^4,28^. Prior to clustering, we standardized the five selected percentiles, ensuring uniform impact and preventing those with higher variability from driving clustering outcomes. Data for cities within each temperature cluster then underwent similar modeling as in the main effects analysis.

For subgroup analyses, we stratified the outcome by binary sex, age, and mode of transportation involved in the fatality. The five age-group categories included: ≤9, 10-19, 20-34, 35-64, and ≥ 65 years, capturing various stages of life with different risk factors due to developmental, behavioral, and mobility differences. Modal stratification included motor-vehicle, motorcycle, bicycle, and pedestrian, using the subcategories per the ICD-10 codes described above.

To analyze city-level effect modification, we incorporated linear interaction terms between daily temperature conditions and the two city-level transportation-related variables described above: mean street-segment length (km) and mean peak-hour travel time (minutes). Interaction effects were assessed using two approaches ^47^. First, Chi-square likelihood ratio tests, conducted as tests of effect modification and based on models reconstructed from the median values of road-traffic mortality across the imputations, evaluated differences between the main effects and interaction models. Second, marginal effects for temperatures evaluated at the 10^th^ and 90^th^ percentiles of the respective city-level variables were plotted to compare how associations varied between cities at the higher and lower ends of each variable’s distribution.

### Sensitivity analyses

Various sensitivity analyses were conducted to evaluate the robustness of our findings. For the main effects analysis, we first used absolute temperature measured in °C as the exposure variable and compared the results to those obtained from a model using relative temperature percentiles. This comparison aimed to assess whether the choice of temperature scale influenced the observed associations, providing insights into the consistency of the exposure-response relationship. Additionally, we tested six alternative knot placements commonly applied in heat-mortality research to evaluate the impact of different spline configurations on model flexibility and fit. These included one knot at the 50th percentile, two knots at percentiles such as 33.3rd and 66.7th, 25th and 75th, and 10th and 90th, as well as three knots at percentiles such as 25th, 50th, and 75th, and 10th, 50th, and 90th ^25^. This analysis was designed to ensure that our conclusions were not overly dependent on a specific choice of knots. Lastly, to complement the stratification analysis by temperature cluster, we also explored effect modification by city-level temperature characteristics, including mean temperature (°C) and temperature standard deviation (°C). These additional analyses aimed to identify whether the observed associations varied across cities with differing climatic profiles, providing deeper insights into how local temperature patterns may influence heat-related risks.

All analyses were performed in R (version 4.3.1). Modeling was conducted using the dlnm (version 2.4.7), gnm (version 1.1.5), and splines (version 4.3.1) packages.

## Acknowledgments

This project was supported by the Wellcome Trust initiative “Our Planet, Our Health” (grant 205177/Z/16/Z). Authors also acknowledge partial support from NIH grant P20MD019221.

## Data availability

Links to the ERA5-Land, WorldPop, and Global Urban Footprint source datasets used to estimate population-weighted ambient temperature as well as final daily temperature outputs are available at https://github.com/Drexel-UHC/salurbal_heat. Vital registration and population data for Brazil, Chile, and Mexico were downloaded from publicly available repositories from statistical agencies in each country. Vital registration and population data for Costa Rica, El Salvador, and Panama were obtained directly from statistical agencies in each country. A link to these agency websites can be accessed via https://drexel.edu/lac/data-evidence/data-acknowledgements/. Urban boundaries and features were obtained from the SALURBAL-Climate project and are freely available at https://data.lacurbanhealth.org/. Please contact the corresponding author about access to any urban features that are not yet published on the SALURBAL-Climate portal. The SALURBAL project welcomes queries from anyone interested in learning more about its dataset and potential access to data. To learn more about SALURBAL’s dataset, visit the SALURBAL project website (https://drexel.edu/lac/salurbal/overview/) or contact the project at. After publication of this study, study protocols, data dictionaries, and requested study data may be made available to interested investigators after they have signed a data use agreement with SALURBAL and if their study proposal, developed in collaboration with SALURBAL investigators, is approved by the SALURBAL proposal and publications committee. Some data may not be available to external investigators because of data confidentiality agreements.

## Code availability

The code repository is available at https://github.com/Drexel-UHC/MS252/tree/submission

## Inclusion and ethics statement

All co-authors are co-investigators and/or trainees in the SALURBAL Group, an international collaboration of researchers from institutions across the Americas. Several co-authors are based at institutions within the study region (i.e., Latin American urban areas). All co-authors have contributed to every stage of the study, from its inception and design to implementation, manuscript development, and the decision to submit for publication.

## Supplementary Information

**Supplementary Fig. 1.**
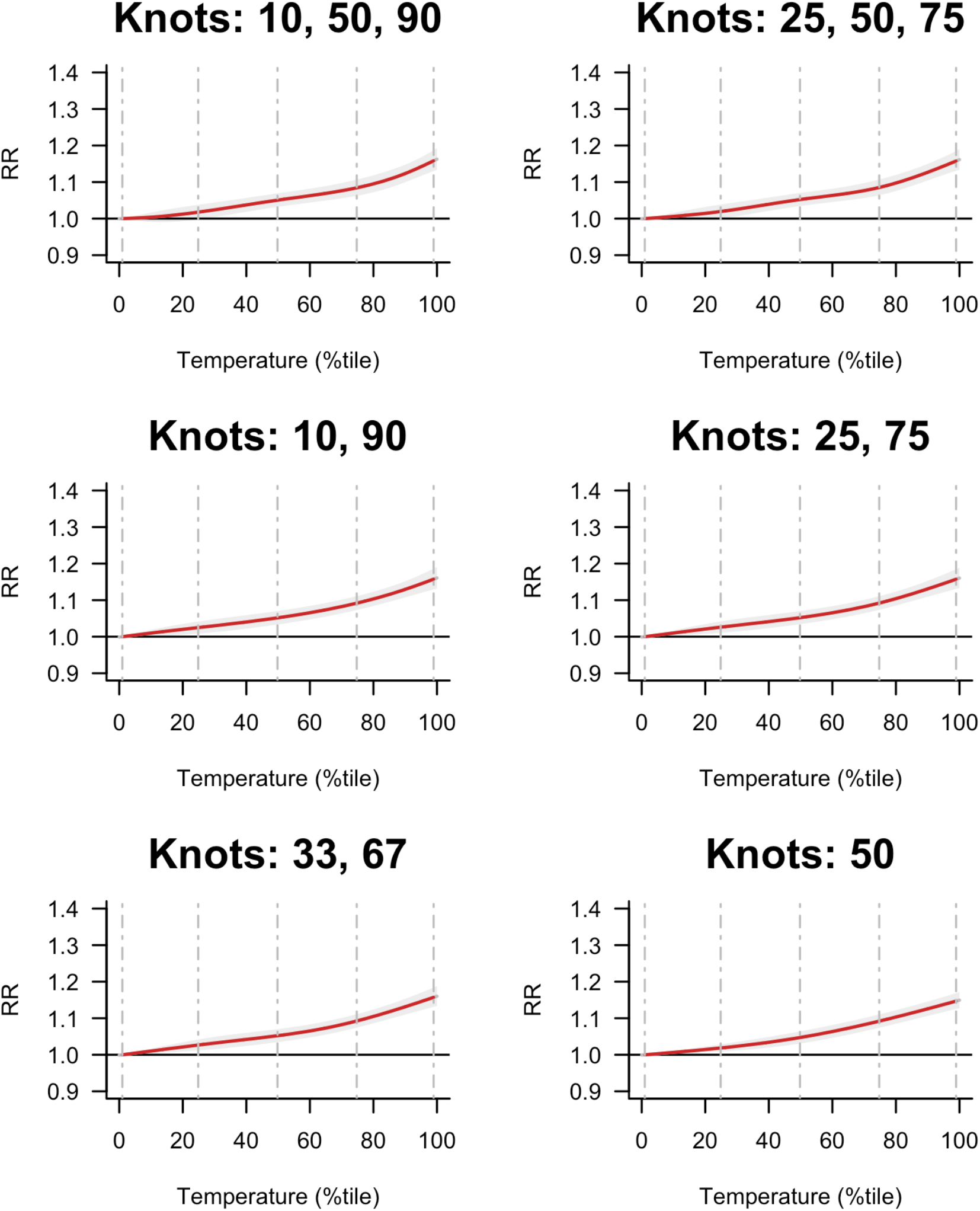
Sensitivity analyses testing different knot specifications. Dashed vertical lines indicate temperature percentiles: 1^st^, 25^th^, 50^th^, 75^th^, and 99^th^ from left to right. Shading in each association curve represents the 95% confidence interval.

**Supplementary Fig. 2.**
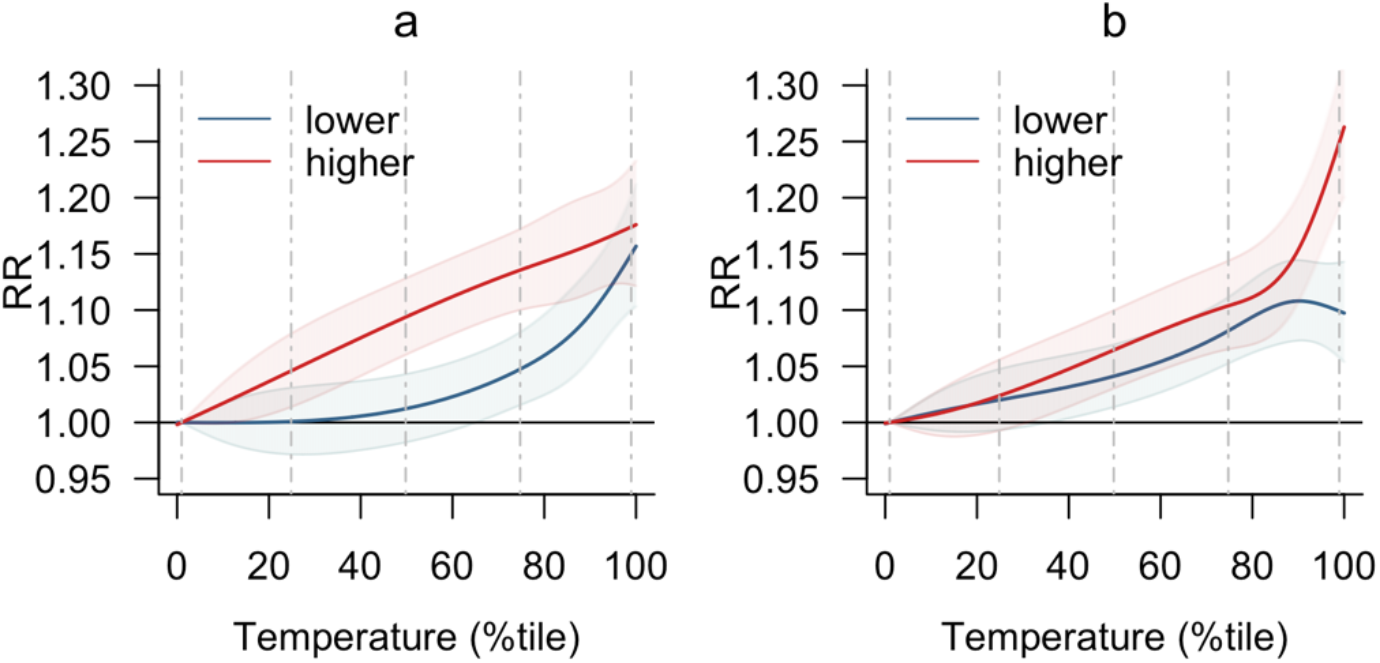
Effect modification by summaries of city-level temperature profiles. (a) Mean temperature. (b) Standard deviation of temperature. High (red) and low (blue) indicate cities at the 90^th^ percentile (higher values) or 10^th^ percentile (lower values) of respective temperature summary. Dashed vertical lines indicate temperature percentiles: 1^st^, 25^th^, 50^th^, 75^th^, and 99^th^ from left to right. Shading in each association curve represents the 95% confidence interval.

## Notes

### Competing Interest Statement

The authors have declared no competing interest.

### Author Declarations

The SALURBAL study protocol was approved by the Drexel University Institutional Review Board (ID no. 1612005035).

